# Effectiveness of Covid-19 vaccines against symptomatic and asymptomatic SARS-CoV-2 infections in an urgent care setting

**DOI:** 10.1101/2022.02.21.22271298

**Authors:** Madhura S. Rane, McKaylee Robertson, Sarah Kulkarni, Daniel Frogel, Chris Gainus, Denis Nash

## Abstract

**Background:** It is critical to monitor changes in vaccine effectiveness against COVID-19 outcomes for various vaccine products in different population subgroups.

**Methods:** We conducted a retrospective study in patients ≥12 years who underwent testing for the SARS-CoV-2 virus from April 1 - October 25, 2021 at urgent care centers in the New York City metropolitan area. Patients self-reported vaccination status at the time of testing. We used a test-negative design to estimate vaccine effectiveness (VE) by comparing odds of a positive test for SARS-CoV-2 infection among vaccinated (n=484,468), partially vaccinated (n=107,573), and unvaccinated (n=466,452) patients, adjusted for demographic factors and calendar time.

**Results:** VE against symptomatic infection after 2 doses of mRNA vaccines was 96% (95% Confidence Interval [CI]: 95%, 97%) in the pre-delta period and reduced to 79% (95% CI: 77%, 81%) in the delta period. In the delta period, VE for 12–15-year-olds (85%; [95% CI: 81%, 89%]) was higher compared to older age groups (<65% for all other age groups). VE estimates did not differ by sex, race/ethnicity, and comorbidity. VE against symptomatic infection was the highest for individuals with a prior infection followed by full vaccination. VE against symptomatic infection after the mRNA-1273 vaccine (83% [95% CI: 81%, 84%]) was higher compared to the BNT162b2 vaccine (76% [95% CI: 74%, 78%]) in the delta period. VE after the single-dose Ad26.COV2.S vaccine was the lowest compared to other vaccines (29% [95% CI: 26%, 32%]) in the delta period.

**Conclusions:** VE against infection after two doses of the mRNA vaccine was high initially, but significantly reduced against the delta variant for all three FDA-approved vaccines.

## INTRODUCTION

As of February 9, 2022, the COVID-19 pandemic has claimed 5.7 million lives globally, with over ∼912,000 deaths reported in the United States (US) ^1^. Vaccines remain the most effective public health tool against COVID-19 morbidity and mortality. In the US, the Pfizer-BioNTech BNT162b2 ^2^ vaccine and the Moderna mRNA-1273 vaccine ^3^ are fully approved and the Janssen Ad26.COV2.S vaccine ^4^ is currently authorized for emergency use. All three vaccines have shown high efficacy against severe disease and mortality in clinical trials against the alpha and delta variants of SARS-CoV-2 virus ^3,5–7^. In November 2021, the highly transmissible B.1.1.529 (Omicron) variant was declared as a variant of concern by the World Health Organization. While evidence of vaccine effectiveness against this variant is still evolving, data from South Africa and UK indicate that mRNA vaccines continue to be effective against serious disease and hospitalization caused the by Omicron variant, particularly among those who were boosted with a third dose ^8,9^.

Observational studies have consistently found high vaccine effectiveness against death and severe disease ^11–17^ for all three vaccines. However, vaccine effectiveness against COVID-19 infection has declined over time, especially since the delta variant emerged as the predominant strain ^11,12,17–24^. As the SARS-CoV-2 virus continues to evolve and more variants emerge ^25^, monitoring changes in vaccine effectiveness against COVID-19 outcomes for various vaccine products in different population subgroups is of utmost importance.

In this study, we used a test-negative case-control design ^26^, comparing vaccination status among people who test positive (cases) and negative (controls), to estimate vaccine effectiveness (VE) against symptomatic and asymptomatic SARS-CoV-2 infection over time in a population of patients seeking care at CityMD, a large ambulatory care center in NYC and neighboring areas. We also compared vaccine-induced and infection-induced protection against SARS-CoV-2 by vaccine product.

## METHODS

### Study Population, Setting, and Design

We conducted a test-negative case control study to estimate vaccine effectiveness of different COVID-19 vaccines among residents of New York City and surrounding metropolitan area who received a polymerase chain reaction (PCR) test or a rapid antigen test for SARS-CoV-2 at one of the 115 CityMD New York locations in the five boroughs of NYC (n=76), Long Island, NY (n=32), and Westchester, NY (n=7).

Individuals ≥ 12 years of age who reported vaccination history and who received at least one PCR or antigen test at CityMD clinics between April 1, 2021, and October 25, 2021 were eligible for the study. We excluded patients <12 years and those with missing vaccination status (3.5%). Only the first positive test for each patient was included. Negative tests performed within 7 days of a previous negative test and within 21 days of a positive test result were excluded, since they could be associated with the same illness ^27^ or be false negatives.

This study was approved by the Institutional Review Board of the City University of New York. Patient consent was not obtained because deidentified electronic health records were used.

### Exposure and outcome measures

#### COVID-19 vaccination

Data on COVID-19 vaccination status was systematically ascertained via patient self-report as part of their intake and history. At the time of testing, patients reported whether they received a COVID-19 vaccine, the vaccine manufacturer, and whether two weeks have elapsed since their final dose. Patients who reported receiving only one dose of the mRNA vaccines or 2 doses within 2 weeks of the day of testing were defined as partially vaccinated. Those receiving 2 doses of the mRNA vaccine at least 2 weeks before testing were defined as fully-vaccinated.

#### SARS-CoV-2 infection - Cases and Controls

Testing for SARS-CoV-2 conducted at CityMD using assays authorized by the Food and Drug Administration (FDA), included: PCR tests of respiratory tract specimens for SARS-CoV-2 RNA collected via nasopharyngeal and nasal swabs and rapid antigen tests of respiratory tract specimens collected via anterior nasal swabs. All patients were evaluated by a licensed clinician.

#### Symptomatic and Asymptomatic SARS-CoV-2 Infection

We defined symptomatic COVID-19 as SARS-CoV-2 positive patients who exhibited at least one of these symptoms: fever, oxygen saturation < 95%, chills, cough, headache, fatigue, myalgia, sore throat, chest tightness, shortness of breath, loss of sense of taste and smell, nausea, vomiting, diarrhea, chest pain, confusion/altered mental state. If no symptoms were recorded in the EMR, patients were considered asymptomatic. Date of visit was used as a proxy for the date of symptom onset.

#### Covariates

Patient age, sex, race/ethnicity, comorbidities, region of residence, and BMI were obtained from the EMR. Individuals with a higher likelihood of COVID-19 exposure are likely to undergo serial testing. Therefore, the total number of tests performed at CityMD before April 1, 2021, were used as proxy for individuals with higher exposure to SARS-CoV-2. We assumed that the absence of information on comorbidities, symptoms, and previous tests in the EMR meant that none were present.

#### Statistical analysis

Chi-squared test was used to compare characteristics between test positive cases and test negative controls and between vaccinated and unvaccinated testers. Multivariable logistic regression was used to estimate the odds ratio (OR) of vaccination comparing cases and controls. VE was calculated as (1-OR) x 100%. ORs were adjusted for the covariates listed above, selected a priori as potential confounders. To account for temporal confounding due to increasing vaccine coverage, different timing of vaccine eligibility for different age groups, and changing SARS-CoV-2 incidence over time, we adjusted the models for calendar time grouped in 2-week intervals ^28^.

### Vaccine effectiveness for mRNA vaccines

For the main analysis, we estimated VE for the 2-dose mRNA vaccines combined. Patients who received the one-dose Jannsen vaccine (6% of the study sample) were excluded. Patients tested positive at any time prior to April 1, 2021, by PCR, rapid test, or antibody tests conducted at CityMD clinics were also excluded. For all models, VE before the delta variant became predominant in the NYC area (April 1, 2021 - June 10, 2021, henceforth referred to as ‘pre-delta period’) and during delta variant predominance (June 11, 2021 - October 25, 2021, ‘delta period’) were compared. VE was also estimated stratified by age group (12-15 years, 16-30 years, 31-50 years, 51-64 years 65-80 years, >80 years), sex, presence of comorbidities, and race/ethnicity.

### Vaccine-induced and infection-induced protection by vaccine product

We compared vaccine-induced and infection-induced protection separately for each vaccine product (Pfizer-BioNTech BNT162b2, Moderna mRNA-1273, and Janssen Ad26.COV2.S), adjusted for the same covariates above. Patients with no vaccine-induced or infection-induced protection (reference group) were compared to patients with a previous infection but no vaccine (infection-induced protection only), previous infection + one or two doses of vaccine (hybrid protection), and one or two doses of vaccine but no previous infection (vaccine-induced protection only).

As a sensitivity analysis to assess whether the type of test used (RT-PCR vs. rapid antigen test) biased VE estimates, we estimated VE for 2-dose mRNA vaccines against any infection restricted to patients who received only the RT-PCR test. All analyses were conducted in R 4.0.1(Vienna, Austria). Tests were two-sided with P-value <0.05 considered significant.

## RESULTS

The study sample included 1,058,493 individuals who contributed at least one COVID-19 RT-PCR or rapid antigen test between April 1, 2021 and October 25, 2021 (Figure 1). The median age of study participants was 33 years (interquartile range, 25 - 49). Patient characteristics by vaccination and SARS-CoV-2 infection status are described in Table 1. A total of 55,827 (5.3%) patients tested positive at least once for the SARS-CoV-2 virus, while 45.8% were fully-vaccinated, 10.1% were partially vaccinated, and 44.1% were unvaccinated at the time of testing. Among positive individuals, 13.6% were symptomatic at the time of testing and 19.2% had at least one comorbidity. The proportion of testers vaccinated at the time of testing increased over time from 29.7% in April 2021 to 71.0% in October 2021 (Supplementary figure 1).

**Table 1:**
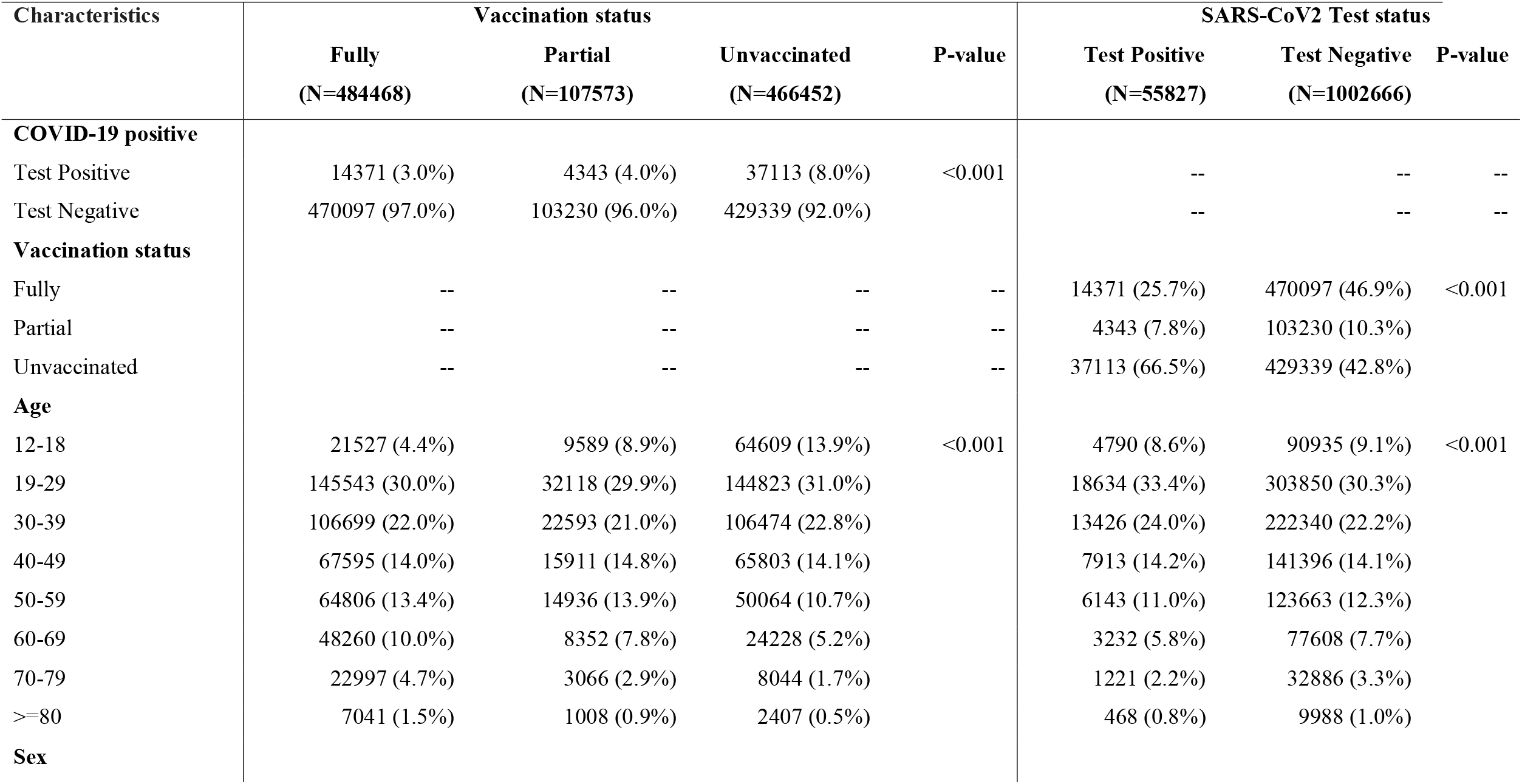
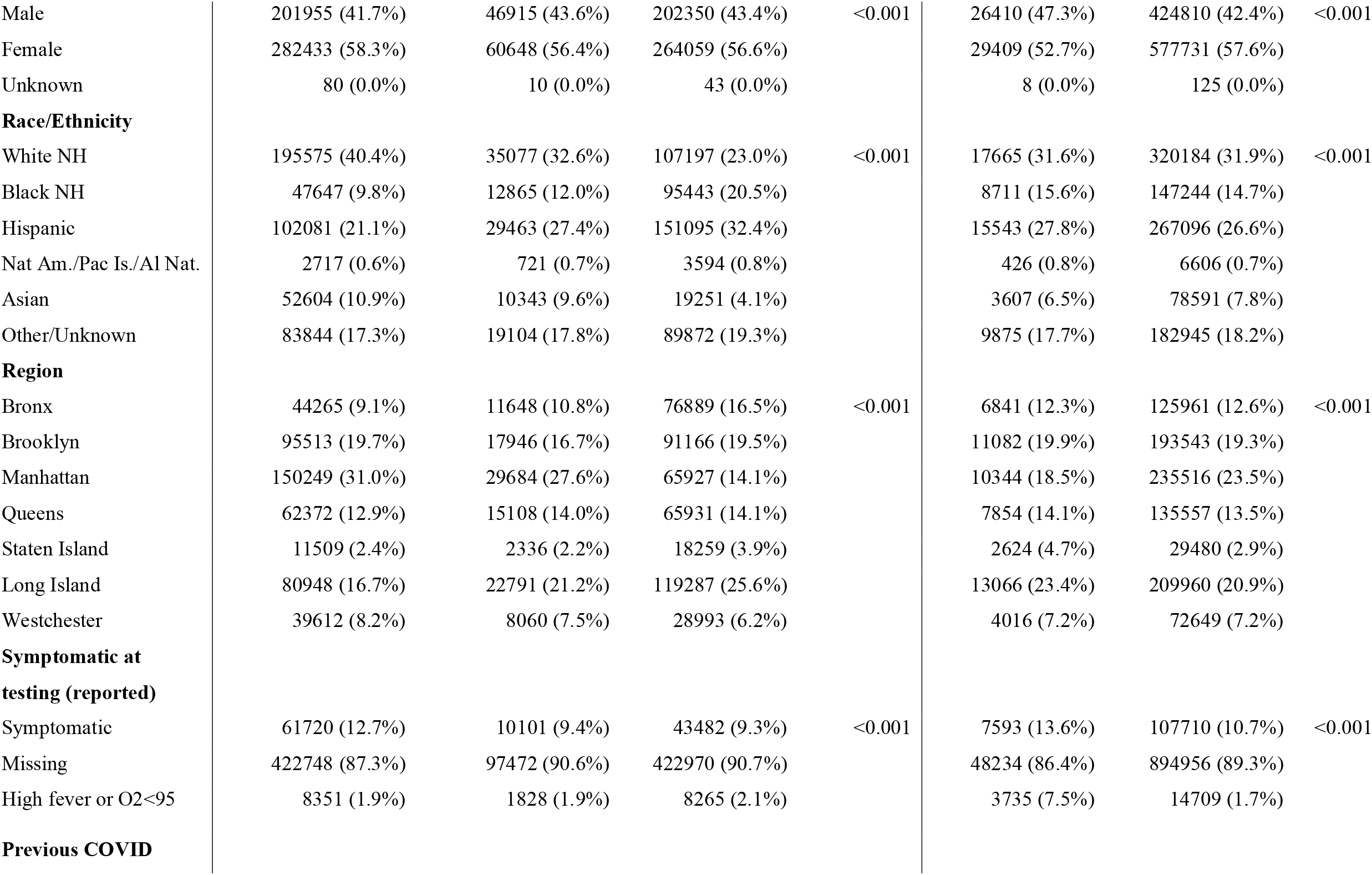
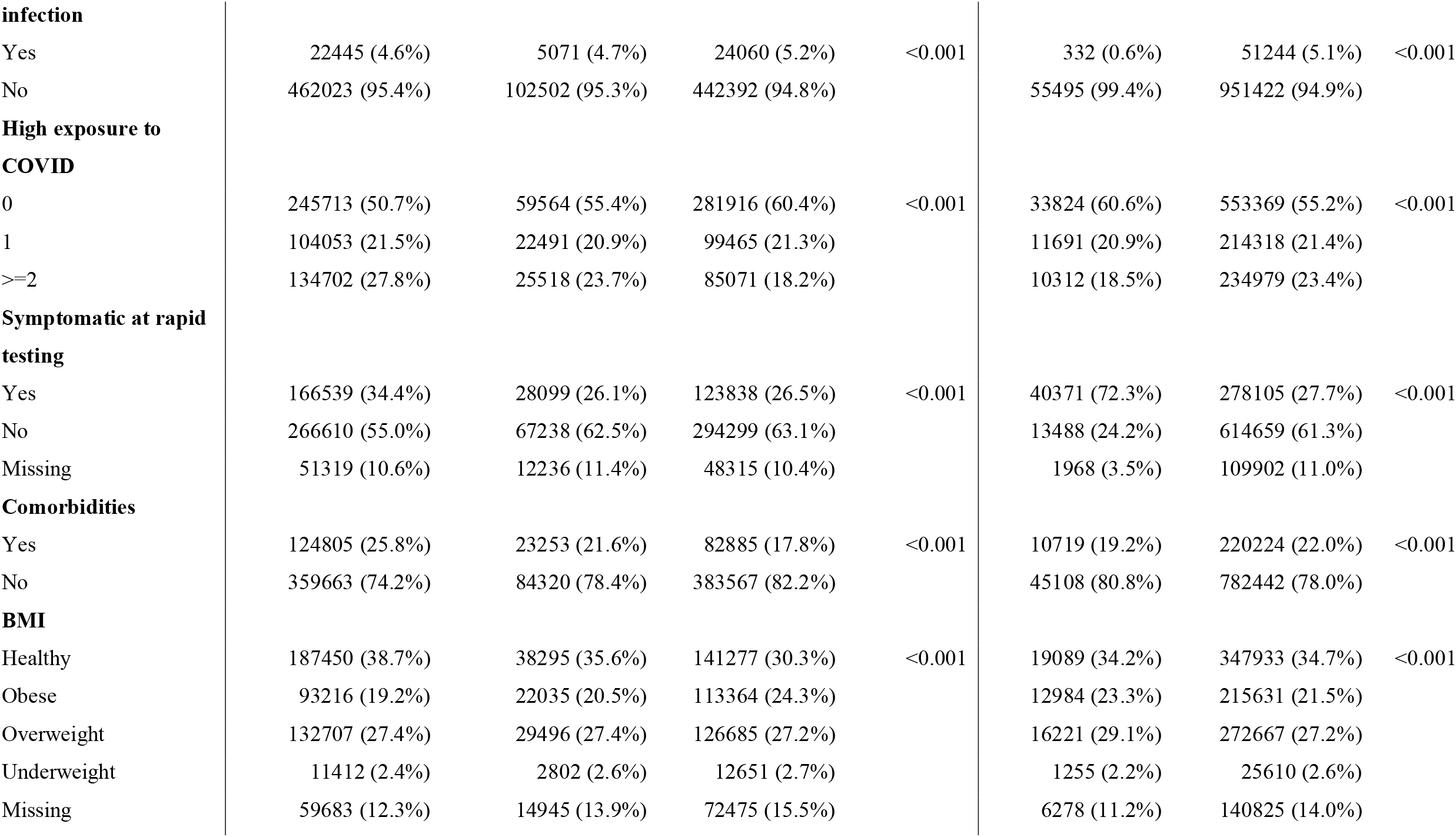
Characteristics of participants tested for SARS-CoV-2 between April 1, 2021, and October 25, 2021, at CityMD clinics in New York City and surrounding metropolitan area

| Characteristics | Vaccination status |  |  |  | SARS-CoV2 Test status |  |  |
| --- | --- | --- | --- | --- | --- | --- | --- |
|  | Fully<br>(N=484468) | Partial<br>(N=107573) | Unvaccinated<br>(N=466452) | P-value | Test Positive<br>(N=55827) | Test Negative<br>(N=1002666) | P-value |
| **COVID-19 positive** |  |  |  |  |  |  |  |
| Test Positive | 14371 (3.0%) | 4343 (4.0%) | 37113 (8.0%) | <0.001 | -- | -- | -- |
| Test Negative | 470097 (97.0%) | 103230 (96.0%) | 429339 (92.0%) |  | -- | -- | -- |
| **Vaccination status** |  |  |  |  |  |  |  |
| Fully | -- | -- | -- | -- | 14371 (25.7%) | 470097 (46.9%) | <0.001 |
| Partial | -- | -- | -- | -- | 4343 (7.8%) | 103230 (10.3%) |  |
| Unvaccinated | -- | -- | -- | -- | 37113 (66.5%) | 429339 (42.8%) |  |
| **Age** |  |  |  |  |  |  |  |
| 12-18 | 21527 (4.4%) | 9589 (8.9%) | 64609 (13.9%) | <0.001 | 4790 (8.6%) | 90935 (9.1%) | <0.001 |
| 19-29 | 145543 (30.0%) | 32118 (29.9%) | 144823 (31.0%) |  | 18634 (33.4%) | 303850 (30.3%) |  |
| 30-39 | 106699 (22.0%) | 22593 (21.0%) | 106474 (22.8%) |  | 13426 (24.0%) | 222340 (22.2%) |  |
| 40-49 | 67595 (14.0%) | 15911 (14.8%) | 65803 (14.1%) |  | 7913 (14.2%) | 141396 (14.1%) |  |
| 50-59 | 64806 (13.4%) | 14936 (13.9%) | 50064 (10.7%) |  | 6143 (11.0%) | 123663 (12.3%) |  |
| 60-69 | 48260 (10.0%) | 8352 (7.8%) | 24228 (5.2%) |  | 3232 (5.8%) | 77608 (7.7%) |  |
| 70-79 | 22997 (4.7%) | 3066 (2.9%) | 8044 (1.7%) |  | 1221 (2.2%) | 32886 (3.3%) |  |
| >=80 | 7041 (1.5%) | 1008 (0.9%) | 2407 (0.5%) |  | 468 (0.8%) | 9988 (1.0%) |  |
| **Sex** |  |  |  |  |  |  |  |
| Male | 201955 (41.7%) | 46915 (43.6%) | 202350 (43.4%) | <0.001 | 26410 (47.3%) | 424810 (42.4%) | <0.001 |
| Female | 282433 (58.3%) | 60648 (56.4%) | 264059 (56.6%) |  | 29409 (52.7%) | 577731 (57.6%) |  |
| Unknown | 80 (0.0%) | 10 (0.0%) | 43 (0.0%) |  | 8 (0.0%) | 125 (0.0%) |  |
| Race/Ethnicity |  |  |  |  |  |  |  |
| White NH | 195575 (40.4%) | 35077 (32.6%) | 107197 (23.0%) | <0.001 | 17665 (31.6%) | 320184 (31.9%) | <0.001 |
| Black NH | 47647 (9.8%) | 12865 (12.0%) | 95443 (20.5%) |  | 8711 (15.6%) | 147244 (14.7%) |  |
| Hispanic | 102081 (21.1%) | 29463 (27.4%) | 151095 (32.4%) |  | 15543 (27.8%) | 267096 (26.6%) |  |
| Nat Am./Pac Is./Al Nat. | 2717 (0.6%) | 721 (0.7%) | 3594 (0.8%) |  | 426 (0.8%) | 6606 (0.7%) |  |
| Asian | 52604 (10.9%) | 10343 (9.6%) | 19251 (4.1%) |  | 3607 (6.5%) | 78591 (7.8%) |  |
| Other/Unknown | 83844 (17.3%) | 19104 (17.8%) | 89872 (19.3%) |  | 9875 (17.7%) | 182945 (18.2%) |  |
| Region |  |  |  |  |  |  |  |
| Bronx | 44265 (9.1%) | 11648 (10.8%) | 76889 (16.5%) | <0.001 | 6841 (12.3%) | 125961 (12.6%) | <0.001 |
| Brooklyn | 95513 (19.7%) | 17946 (16.7%) | 91166 (19.5%) |  | 11082 (19.9%) | 193543 (19.3%) |  |
| Manhattan | 150249 (31.0%) | 29684 (27.6%) | 65927 (14.1%) |  | 10344 (18.5%) | 235516 (23.5%) |  |
| Queens | 62372 (12.9%) | 15108 (14.0%) | 65931 (14.1%) |  | 7854 (14.1%) | 135557 (13.5%) |  |
| Staten Island | 11509 (2.4%) | 2336 (2.2%) | 18259 (3.9%) |  | 2624 (4.7%) | 29480 (2.9%) |  |
| Long Island | 80948 (16.7%) | 22791 (21.2%) | 119287 (25.6%) |  | 13066 (23.4%) | 209960 (20.9%) |  |
| Westchester | 39612 (8.2%) | 8060 (7.5%) | 28993 (6.2%) |  | 4016 (7.2%) | 72649 (7.2%) |  |
| Symptomatic at testing (reported) |  |  |  |  |  |  |  |
| Symptomatic | 61720 (12.7%) | 10101 (9.4%) | 43482 (9.3%) | <0.001 | 7593 (13.6%) | 107710 (10.7%) | <0.001 |
| Missing | 422748 (87.3%) | 97472 (90.6%) | 422970 (90.7%) |  | 48234 (86.4%) | 894956 (89.3%) |  |
| High fever or O2<95 | 8351 (1.9%) | 1828 (1.9%) | 8265 (2.1%) |  | 3735 (7.5%) | 14709 (1.7%) |  |
| Previous COVID |  |  |  |  |  |  |  |
| **infection** |  |  |  |  |  |  |  |
| Yes | 22445 (4.6%) | 5071 (4.7%) | 24060 (5.2%) | <0.001 | 332 (0.6%) | 51244 (5.1%) | <0.001 |
| No | 462023 (95.4%) | 102502 (95.3%) | 442392 (94.8%) |  | 55495 (99.4%) | 951422 (94.9%) |  |
| **High exposure to COVID** |  |  |  |  |  |  |  |
| 0 | 245713 (50.7%) | 59564 (55.4%) | 281916 (60.4%) | <0.001 | 33824 (60.6%) | 553369 (55.2%) | <0.001 |
| 1 | 104053 (21.5%) | 22491 (20.9%) | 99465 (21.3%) |  | 11691 (20.9%) | 214318 (21.4%) |  |
| >=2 | 134702 (27.8%) | 25518 (23.7%) | 85071 (18.2%) |  | 10312 (18.5%) | 234979 (23.4%) |  |
| **Symptomatic at rapid testing** |  |  |  |  |  |  |  |
| Yes | 166539 (34.4%) | 28099 (26.1%) | 123838 (26.5%) | <0.001 | 40371 (72.3%) | 278105 (27.7%) | <0.001 |
| No | 266610 (55.0%) | 67238 (62.5%) | 294299 (63.1%) |  | 13488 (24.2%) | 614659 (61.3%) |  |
| Missing | 51319 (10.6%) | 12236 (11.4%) | 48315 (10.4%) |  | 1968 (3.5%) | 109902 (11.0%) |  |
| **Comorbidities** |  |  |  |  |  |  |  |
| Yes | 124805 (25.8%) | 23253 (21.6%) | 82885 (17.8%) | <0.001 | 10719 (19.2%) | 220224 (22.0%) | <0.001 |
| No | 359663 (74.2%) | 84320 (78.4%) | 383567 (82.2%) |  | 45108 (80.8%) | 782442 (78.0%) |  |
| **BMI** |  |  |  |  |  |  |  |
| Healthy | 187450 (38.7%) | 38295 (35.6%) | 141277 (30.3%) | <0.001 | 19089 (34.2%) | 347933 (34.7%) | <0.001 |
| Obese | 93216 (19.2%) | 22035 (20.5%) | 113364 (24.3%) |  | 12984 (23.3%) | 215631 (21.5%) |  |
| Overweight | 132707 (27.4%) | 29496 (27.4%) | 126685 (27.2%) |  | 16221 (29.1%) | 272667 (27.2%) |  |
| Underweight | 11412 (2.4%) | 2802 (2.6%) | 12651 (2.7%) |  | 1255 (2.2%) | 25610 (2.6%) |  |
| Missing | 59683 (12.3%) | 14945 (13.9%) | 72475 (15.5%) |  | 6278 (11.2%) | 140825 (14.0%) |  |

**Figure 1:**
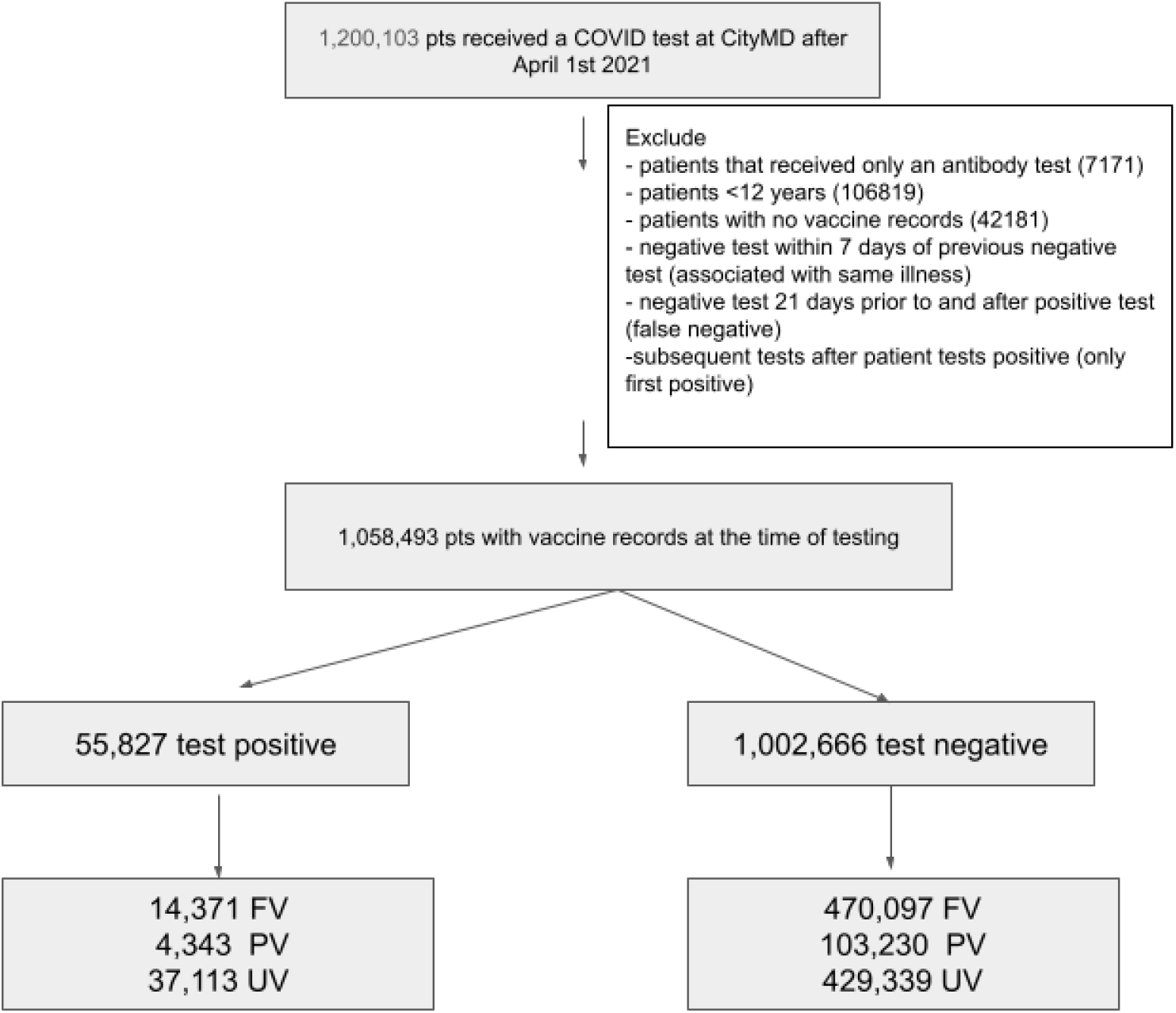
Flow chart of study participants tested for SARS-CoV-2 between April 1 2021 and October 25 2021 at CityMD clinics in New York City and surrounding metropolitan area FV: Fully vaccinated; PV: Partially vaccinated; UV: Unvaccinated

### Vaccine effectiveness over time

Vaccine effectiveness (VE) against any SARS-CoV-2 infection (symptomatic or asymptomatic) after 2 doses of mRNA vaccines (BioNTech BNT162b and mRNA-1273 combined) decreased with calendar time. Adjusted VE after 2 doses of mRNA vaccines was high (88% [95% CI: 87%, 90%]) in April 2021. By mid-June 2021, VE dropped to 69% (95% CI: 62%, 74%), and by October 2021, declined further to 51% (95% CI: 49%, 53%) (Figure 2).

**Figure 2:**
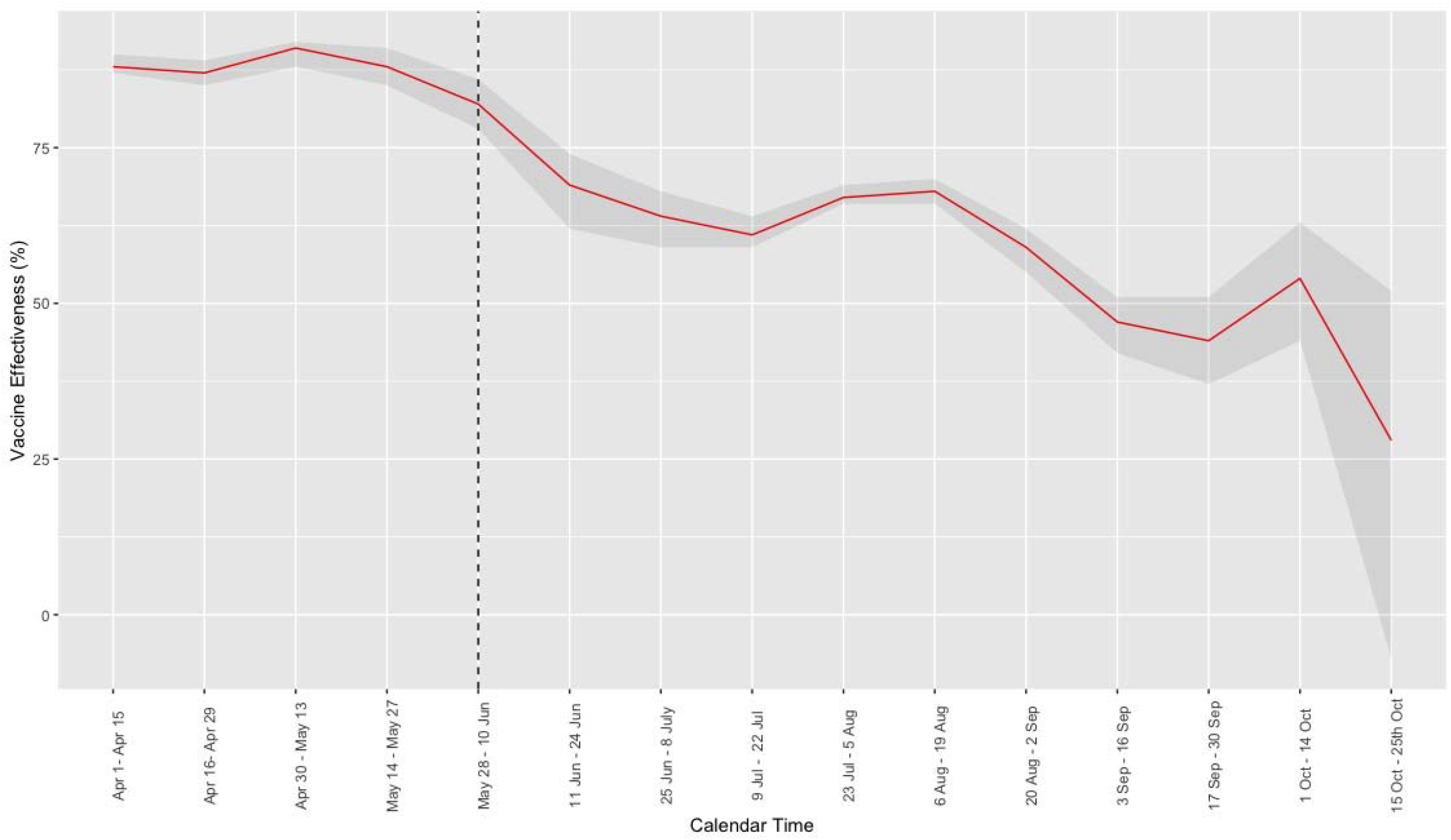
Time-varying vaccine effectiveness against symptomatic and asymptomatic SARS-CoV-2 infections after 2-dose mRNA vaccines for patients testing in CityMD clinics between April 1, 2021 - October 25, 2021 The red line indicates vaccine effectiveness estimates for every 2-week interval during the study period and the 95% CI are shaded in grey. Both symptomatic and asymptomatic infections were included. The dashed horizontal line indicates the start of Delta period.

### Vaccine effectiveness by variant period and subgroups

Overall, adjusted VE with 2-dose mRNA vaccines against symptomatic infection was 96% (95% CI: 95%, 97%) in the pre-delta period and reduced to 79% (95% CI: 77%, 81%) in the delta period. When symptoms were restricted to high fever (>101.4 deg) or O_2_<95%, VE against symptomatic infection increased slightly to 84% (95% CI: 81%, 85%) in the delta period. VE against asymptomatic infection was lower than symptomatic infection (78% [95% CI: 77%, 80%] in the pre-delta period and 59% [95% CI: 58%, 60%]) in the delta period) (Figure 3).

**Figure 3:**
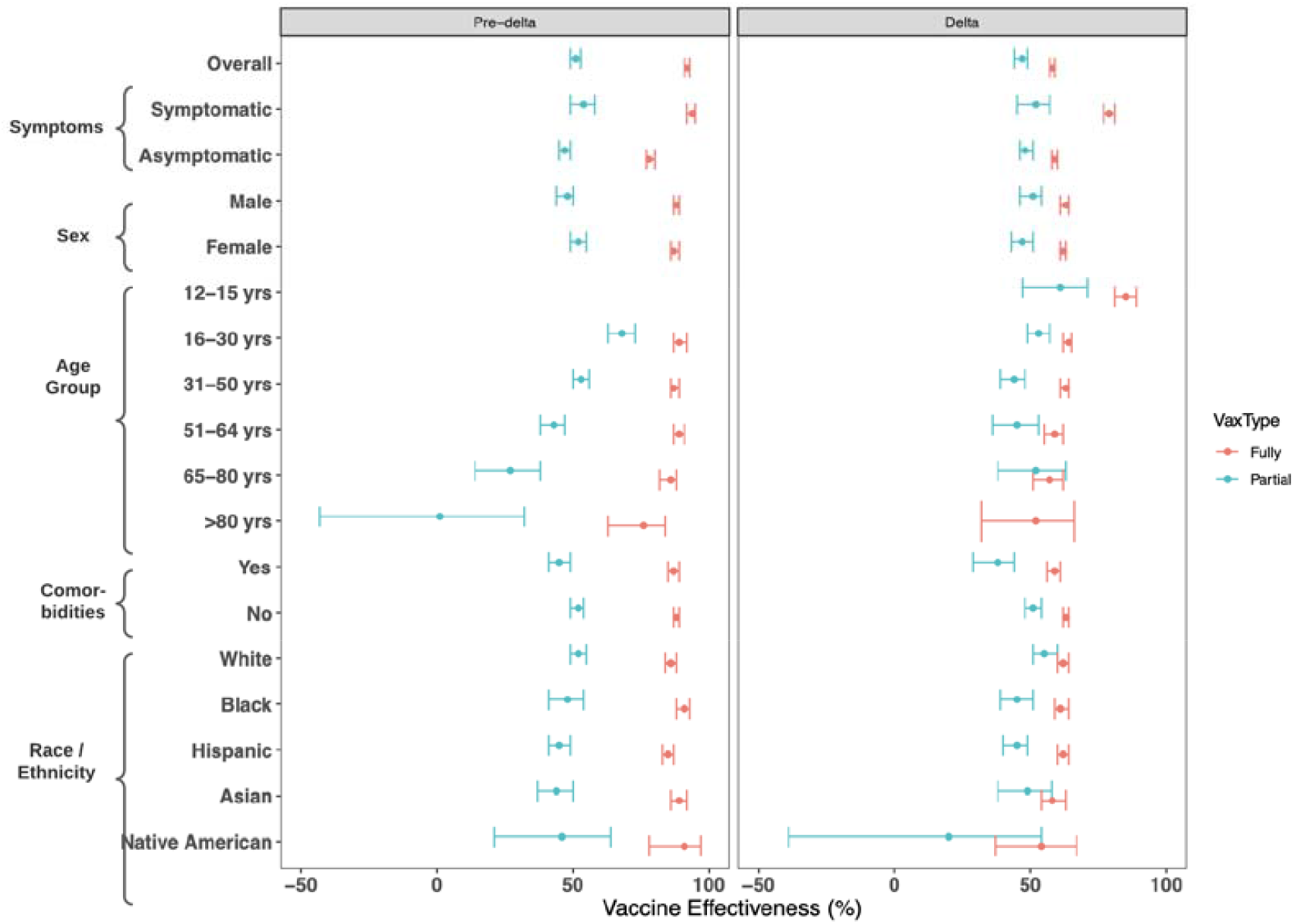
Estimated vaccine effectiveness after 2-dose mRNA vaccines against symptomatic and asymptomatic SARS-CoV-2 infection in different population subgroups mRNA vaccine effectiveness and 95% confidence intervals after 2 doses (red) and 1 dose (blue) of the vaccine for different population subgroups in pre-delta (left panel) and delta (right panel) periods are displayed here. Both symptomatic and asymptomatic SARS-CoV-2 cases were included for the overall VE estimates and the VE estimates by subgroups. Patients 12-15 years old became eligible for COVID-19 vaccination on May 12, 2021 so vaccination information for this age group was available for the pre-delta period. Similarly, there were no patients >80 years of age who had received one 1 dose of an mRNA vaccine in the delta period.

Adjusted VE with 2-dose mRNA vaccines against any infection was higher in the pre-delta compared to delta period for all demographic subgroups. VE for ages 16 to 64 years was >85% in the pre-delta period vs. <65% in delta period. VE was lower for 64+ year olds compared with <64-year-olds, even in the pre-delta period. Compared to age groups older than 16 years, higher VE was observed for 12– 15-year-olds (85%; [95% CI: 81%, 89%]) in the delta period. VE estimates were comparable across sex, race/ethnicity, and comorbidity groups in both pre-delta and delta period (Figure 3).

### Comparison between vaccine-induced and infection-induced protection, by vaccine product

A total of 51,576 participants had previous infections documented between March 1, 2020 and March 31, 2021 (average time between prior infection and second infection during study period: 236 days, range: 46 - 587). Protection against symptomatic infection was the highest for those fully-vaccinated by any of the vaccines and had a history of infection (Figure 4a, 4b, 4c). Compared to those unvaccinated with no prior infections, those unvaccinated **with** prior infections had a 91% (95% CI: 86%, 94%) reduction in odds of symptomatic infection in the pre-delta period and 89% (95% CI: 83%, 93%) reduction in odds in the delta period (Figure 4).

**Figure 4:**
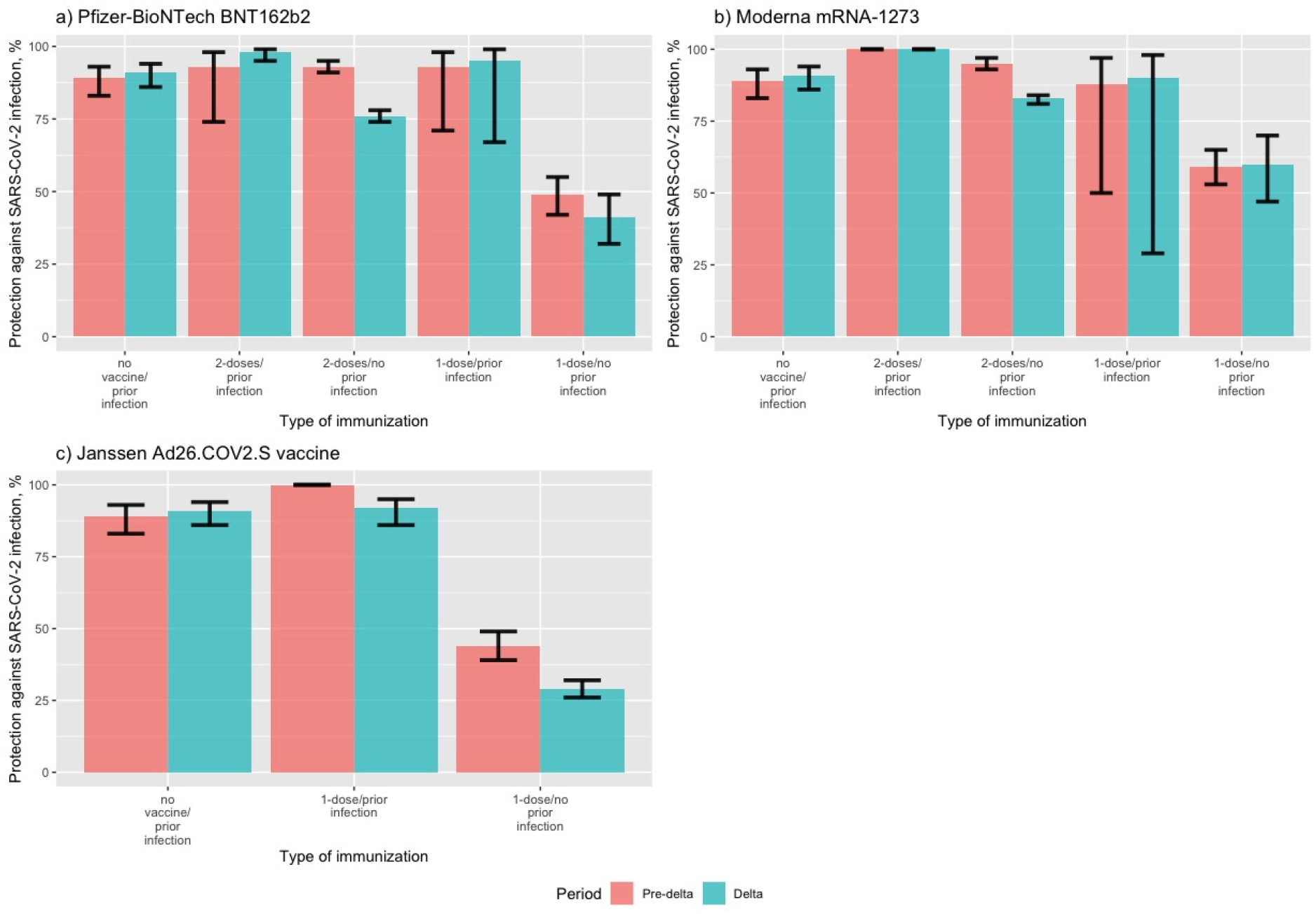
Estimated vaccine-induced and infection-induced protection against *<u>symptomatic</u>* SARS-CoV-2 infection by vaccine product Bar graph shows protection and 95% confidence intervals against SARS-CoV-2 symptomatic infection comparing patients with infection-induced protection, vaccine-induced protection, or both, to patients who were unvaccinated (no vaccine-induced protection) and had no prior infections (no infection-induced protection). Results are shown separately for the (a) Pfizer-BioNTech BNT 162b2 vaccine, (b) Moderna mRNA-1273 vaccine, and (c) Janssen Ad26.COV2.S vaccine. Protection during pre-delta period is in red while protection during delta period is in blue.

Among those **without** documented prior infection, VE against symptomatic infection after 2 doses of the BNT162b2 vaccine was 93% (95% CI: 90%, 95%) in the pre-delta period and declined to 76% (95% CI: 74%, 78%) in the delta period (Figure 4a). VE after 2 doses of the mRNA-1273 vaccine against symptomatic infection was higher compared to the BNT162b2 vaccine, particularly in the delta period (95% [95% CI: 93%, 97%] in the pre-delta period and 83% [95% CI: 81%, 84%] in the delta period) (Figure 4c). VE after 1 dose of the Ad26.COV2.S vaccine was much lower compared to the 2-dose mRNA vaccines (44% [95% CI: 39%, 49%] in the pre-delta period and 29% [95% CI: 26%, 32%] in the delta period) (Figure 4c). Similar trends were observed for VE against asymptomatic infections (Figure 5).

**Figure 5:**
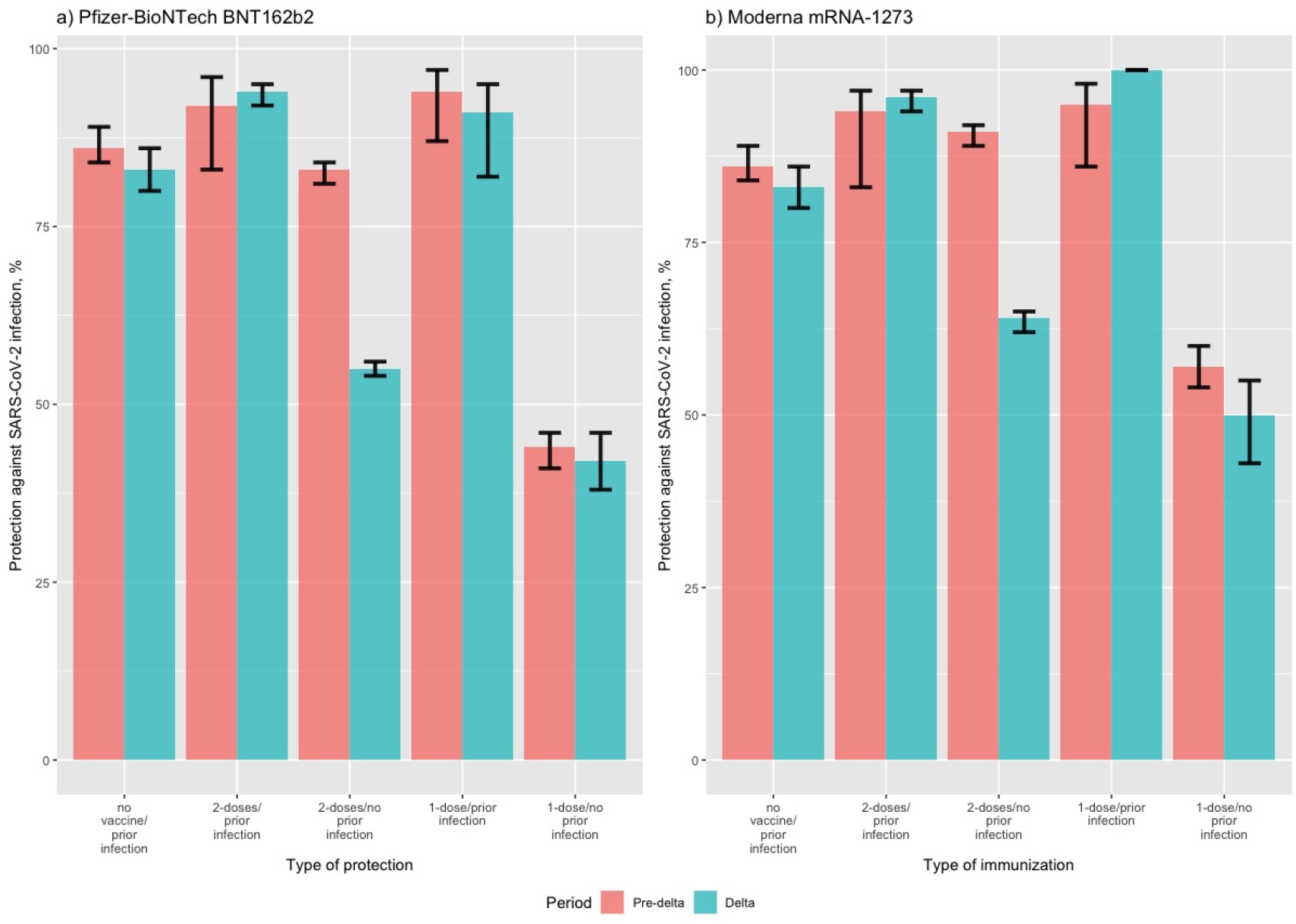
Estimated vaccine-induced and infection-induced protection against *<u>asymptomatic</u>* SARS-CoV-2 infection by vaccine product Bar graph shows protection and 95% confidence intervals against SARS-CoV-2 asymptomatic infection comparing patients with infection-induced protection, vaccine-induced protection, or both, to patients who were unvaccinated (no vaccine-induced protection) and had no prior infections (no infection-induced protection). Results are shown separately for the (a) Pfizer-BioNTech BNT 162b2 vaccine, and (b) Moderna mRNA-1273 vaccine. No patients who received the Janssen Ad26.COV2.S vaccine reported an asymptomatic infection. Protection during pre-delta period is in red while protection during delta period is in blue.

In sensitivity analyses, VE estimates against any SARS-CoV-2 infection detected by RT-PCR only and by both RT-PCR and antigen tests were consistent for both the 2-dose mRNA vaccines and the 1-dose Ad26.COV2.S vaccine (<u>Supplementary Table 1</u>).

## Discussion

Using a test-negative case-control study design, we estimated vaccine effectiveness against symptomatic and asymptomatic SARS-CoV2 infection in a population of testers at urgent care centers in the NYC metropolitan area during the pre-delta and delta eras of the pandemic. We found that vaccine effectiveness after 2 doses of mRNA vaccines was very high (>90%) in the pre-delta period across subgroups of populations but declined over time. The mRNA-1273 vaccine was found to be slightly more effective compared to the BNT162b2 vaccine, while VE was much lower for recipients of the Ad26.COV2.S vaccine. Individuals with both prior infection and vaccination had the highest protection against re-infection. Comparisons between effectiveness of different vaccine products and between infection-induced, vaccine-induced, and hybrid protection can help inform policy decisions about boosters.

Because the study participants are restricted to those who attend an urgent care clinic for a test, the test-negative study design can potentially control for selection bias arising due to differences in healthcare-seeking behavior between cases and controls. Moreover, use of highly specific and sensitive molecular tests for case detection makes outcome misclassification less likely. ^26^. This design has been popular for estimating COVID-19 vaccine effectiveness ^12,23,29,30^. Our study contributes to a broader understanding of real-world COVID-19 vaccine effectiveness in key demographic groups. First, we estimated VE within population subgroups disproportionately affected by COVID-19, such as individuals >80 years, adolescents, Black and Hispanic adults, and those with comorbidities. Second, we were able to compare VE for all three FDA-approved and authorized vaccines in the US. Lastly, we compared infection-induced and vaccine-induced protection against both symptomatic and asymptomatic infections, addressing a key gap in COVID-19 vaccine effectiveness research.

Our estimates for VE against symptomatic infection for both partial and full vaccination with mRNA vaccines are comparable to findings of other studies performed in different settings and populations, especially for the pre-delta period ^30–35^. Comparing the two mRNA vaccine products, we found higher VE estimates for the mRNA-1273 vaccine consistent with other studies ^16,36,37^. The difference in effectiveness between the two vaccines might be because of higher antigen dose in the mRNA-1273 vaccine or longer interval between doses ^37,38^. The VE estimates for the Ad26.COV2.S vaccine were much lower compared to other published reports ^19,29,39^, particularly in the delta period. The reason for lower VE for the Ad26.COV2.S vaccine in general might be lower yields of antibody titres among Ad26.COV2.S vaccine recipients compared to mRNA vaccine recipients ^40^.

The stark drop in VE in early July is contemporaneous with the rise in prevalence of the Delta variant in New York City, which accounted for 60% of all cases by the end of June ^41^. Some studies posit that decrease in VE is likely to be due to lower effectiveness against the delta variant ^5,39^, while others have attributed it to waning of vaccine effect ^11,32,42^. As we did not have vaccination dates in our study, we could not directly differentiate between these two mechanisms. However, higher delta-period VE estimates for 12–15-year-olds (who were vaccinated more recently) compared to VE estimates for 65+ year olds (who were vaccinated in early 2021) could suggest a possible waning effect. While data is still emerging, mRNA vaccine effectiveness against infection with the Omicron variant might be too low to prevent outbreaks and super spreading at current vaccination coverage levels in the US. Given these uncertainties, it remains important to monitor how VE changes over time especially as new variants continue to emerge ^43^.

VE against infection among >65-year-olds, as well as among those with comorbidities, had declined to under 60% by October 2021. These findings support CDC’s recommendations to prioritize booster doses for these individuals, given their elevated risk of mortality and hospitalization ^44 45^. VE was comparable across racial/ethnic groups, which is an important finding given the high burden of disease and the initially low COVID-19 vaccine acceptance in racial minorities in the US ^46,47^. Recently, gaps in vaccination coverage by race/ethnicity have narrowed, with a majority of unvaccinated people in the US now being White ^48^. Reporting vaccine effectiveness estimates by race/ethnicity can help to increase vaccine confidence among communities with low vaccine acceptance.

VE estimates comparing vaccine-induced and infection-induced protection from (re)-infection is scarce, complicated by the timing of vaccine roll-out in most populations less than a year into the pandemic. In line with other studies, we found that, in the pre-delta period, vaccine-induced and infection-induced protection were comparable for both mRNA vaccines, and vaccination following infection provided incremental protection against COVID-19 ^49–51^. In the delta period, however, a history of infection was found to be more protective against re-infections compared to vaccination without prior infection, similar to evidence found in Israel and India ^52,53^, but in contrast to evidence from the US ^54^. In our study, the relatively short average time since previous infection (7 months) could be the reason for higher protection among those with prior infection ^51,55–57^.

Individuals with previous infection and one dose of the either mRNA vaccine had high protection against re-infection, suggesting that the first dose of the mRNA vaccine may act like a booster for those who were previously infected ^58^. Even so, full vaccination is still highly recommended for those with prior infection, because levels of protection can vary based on age and disease severity ^59,60^, while VE is relatively uniform across populations, with the possible exception of older and immunocompromised individuals ^61^. Furthermore, even mild disease can lead to long COVID-19 ^62,63^.

Our study has limitations. Even though test-negative design controls for bias due to healthcare-seeking behavior and access, the rates of testing between vaccinated and unvaccinated people might differ. Vaccinated individuals with symptoms could be more motivated to get tested which would lead to an overrepresentation of vaccinated individuals with positive test results, biasing the VE estimates downwards ^64^. Bias due to unmeasured confounders, such as occupation or behavior changes following policies or vaccination is also possible ^65,66^. We may have missed reports of prior infections if participants were tested at other clinics, which could also bias VE estimates downwards. Outcomes for patients who received only an antigen test could be misclassified due to lower test sensitivity. However, our sensitivity analysis showed that when restricted to the highly sensitive RT-PCR tests results, VE estimates were nearly identical to those of the overall sample. Misclassification of exposure is a possibility as vaccination status was self-reported and dates of vaccination were not collected. However, other studies have shown very high agreement between self-reported and recorded COVID-19 vaccination status ^67,68^. VE after one dose of mRNA vaccines is likely underestimated in this study because it was not known if the outcome occurred more than 15 days after the first dose. We did not collect information on disease severity and hospitalization and could not differentiate between asymptomatic and pre-symptomatic patients. These results are not generalizable to populations that do not seek healthcare at ambulatory clinics in NYC.

In conclusion, our analysis of urgent care visit data over a 7-month period showed high overall effectiveness after two doses of the mRNA vaccine in the pre-delta period, but significantly reduced vaccine effectiveness in the delta period for all three FDA-approved vaccines. Prior infection combined with full vaccination provided highest protection regardless of which variant was predominant. Our findings support continued monitoring of vaccine effectiveness by vaccine product as new variants emerge.

## Data Availability

Data may be available upon reasonable request submitted to CityMD/ Summit Health due to privacy/ethical restrictions in using electronic medical records.

## References

1. Center for Systems Science and Engineering (CSSE) at Johns Hopkins University (JHU). COVID-19 Dashboard by the Center for Systems Science and Engineering (CSSE) at Johns Hopkins University (JHU). https://coronavirus.jhu.edu/ (2021).

2. Polack, F. P. et al. Safety and Efficacy of the BNT162b2 mRNA Covid-19 Vaccine. N. Engl. J. Med. 383, 2603–2615 (2020).

3. Baden, L. R. et al. Efficacy and Safety of the mRNA-1273 SARS-CoV-2 Vaccine. N. Engl. J. Med. 384, 403–416 (2021).

4. Sadoff, J. et al. Safety and Efficacy of Single-Dose Ad26.COV2.S Vaccine against Covid-19. N. Engl. J. Med. 384, 2187–2201 (2021).

5. Thomas, S. J. et al. Safety and Efficacy of the BNT162b2 mRNA Covid-19 Vaccine through 6 Months. N. Engl. J. Med. 385, 1761–1773 (2021).

6. Stephenson, K. E. et al. Immunogenicity of the Ad26.COV2.S Vaccine for COVID-19. JAMA 325, 1535–1544 (2021).

7. Frenck, R. W., Jr et al. Safety, Immunogenicity, and Efficacy of the BNT162b2 Covid-19 Vaccine in Adolescents. N. Engl. J. Med. 385, 239–250 (2021).

8. Collie, S., Champion, J., Moultrie, H., Bekker, L.-G. & Gray, G. Effectiveness of BNT162b2 Vaccine against Omicron Variant in South Africa. N. Engl. J. Med. (2021) doi:10.1056/NEJMc2119270.

9. UK Health Security Agency. SARS-CoV-2 variants of concern and variants under investigation in England Technical briefing: Update on hospitalisation and vaccine effectiveness for Omicron VOC-21NOV-01 (B.1.1.529). (2021).

10. Halloran, M. E. & Hudgens, M. G. Estimating population effects of vaccination using large, routinely collected data. Stat. Med. (2017) doi:10.1002/sim.7392.

11. Lin, D. et al. Effectiveness of Covid-19 vaccines in the United States over 9 months: Surveillance data from the state of North Carolina. bioRxiv (2021) doi:10.1101/2021.10.25.21265304.

12. Ranzani, O. T. et al. Vaccine effectiveness of Ad26.COV2.S against symptomatic COVID-19 and clinical outcomes in Brazil: a test-negative study design. bioRxiv (2021) doi:10.1101/2021.10.15.21265006.

13. Saciuk, Y. et al. Pfizer-BioNTech Vaccine Effectiveness Against SARS-CoV-2 Infection: Findings From a Large Observational Study in Israel. SSRN Electronic Journal doi:10.2139/ssrn.3868853.

14. Glatman-Freedman, A., Bromberg, M., Dichtiar, R., Hershkovitz, Y. & Keinan-Boker, L. The BNT162b2 vaccine effectiveness against new COVID-19 cases and complications of breakthrough cases: A nation-wide retrospective longitudinal multiple cohort analysis using individualised data. EBioMedicine vol. 72 103574 (2021).

15. Xu, S. et al. COVID-19 Vaccination and Non--COVID-19 Mortality Risk—Seven Integrated Health Care Organizations, United States, December 14, 2020--July 31, 2021. MMWR Surveill. Summ. 70, 1520 (2021).

16. Puranik, A. et al. Comparison of Two Highly-Effective mRNA Vaccines for COVID-19 During Periods of Alpha and Delta Variant Prevalence. SSRN Electronic Journal doi:10.2139/ssrn.3902782.

17. Bruxvoort, K. J. et al. Real-world effectiveness of the mRNA-1273 vaccine against COVID-19: Interim results from a prospective observational cohort study. Lancet Reg Health Am 100134 (2021).

18. Goldberg, Y. et al. Waning Immunity after the BNT162b2 Vaccine in Israel. N. Engl. J. Med. (2021) doi:10.1056/NEJMoa2114228.

19. Corchado-Garcia, J. et al. Analysis of the Effectiveness of the Ad26.COV2.S Adenoviral Vector Vaccine for Preventing COVID-19. JAMA Netw Open 4, e2132540 (2021).

20. Nordström, P., Ballin, M. & Nordström, A. Effectiveness of Covid-19 Vaccination Against Risk of Symptomatic Infection, Hospitalization, and Death Up to 9 Months: A Swedish Total-Population Cohort Study. (2021) doi:10.2139/ssrn.3949410.

21. Emborg, H.-D. et al. Vaccine effectiveness of the BNT162b2 mRNA COVID-19 vaccine against RT-PCR confirmed SARS-CoV-2 infections, hospitalisations and mortality in prioritised risk groups. doi:10.1101/2021.05.27.21257583.

22. Regev-Yochay, G. et al. Decreased infectivity following BNT162b2 vaccination: A prospective cohort study in Israel. Lancet Reg Health Eur 7, 100150 (2021).

23. Tang, P. et al. BNT162b2 and mRNA-1273 COVID-19 vaccine effectiveness against the SARS-CoV-2 Delta variant in Qatar. Nat. Med. (2021) doi:10.1038/s41591-021-01583-4.

24. Bruxvoort, K. J. et al. Effectiveness of mRNA-1273 against Delta, Mu, and other emerging variants. bioRxiv (2021) doi:10.1101/2021.09.29.21264199.

25. National Center for Immunization and Respiratory Diseases, (NCIRD), Division of Viral Diseases, Centers for Disease Control and Prevention. Science Brief: Omicron (B.1.1.529) Variant. https://www.cdc.gov/coronavirus/2019-ncov/science/science-briefs/scientific-brief-omicron-variant.html (2021).

26. Jackson, M. L. & Nelson, J. C. The test-negative design for estimating influenza vaccine effectiveness. Vaccine 31, 2165–2168 (2013).

27. van Kampen, J. J. A. et al. Duration and key determinants of infectious virus shedding in hospitalized patients with coronavirus disease-2019 (COVID-19). Nat. Commun. 12, 267 (2021).

28. Dean, N. E., Halloran, M. E. & Longini, I. M., Jr. Temporal Confounding in the Test-Negative Design. Am. J. Epidemiol. 189, 1402–1407 (2020).

29. Thompson, M. G. et al. Effectiveness of Covid-19 Vaccines in Ambulatory and Inpatient Care Settings. N. Engl. J. Med. 385, 1355–1371 (2021).

30. Butt, A. A., Omer, S. B., Yan, P., Shaikh, O. S. & Mayr, F. B. SARS-CoV-2 Vaccine Effectiveness in a High-Risk National Population in a Real-World Setting. Ann. Intern. Med. 174, 1404–1408 (2021).

31. Chung, H. et al. Effectiveness of BNT162b2 and mRNA-1273 covid-19 vaccines against symptomatic SARS-CoV-2 infection and severe covid-19 outcomes in Ontario, Canada: test negative design study. BMJ n1943 (2021) doi:10.1136/bmj.n1943.

32. Tartof, S. Y. et al. Effectiveness of mRNA BNT162b2 COVID-19 vaccine up to 6 months in a large integrated health system in the USA: a retrospective cohort study. Lancet 398, 1407–1416 (2021).

33. Thompson, M. G. et al. Interim Estimates of Vaccine Effectiveness of BNT162b2 and mRNA-1273 COVID-19 Vaccines in Preventing SARS-CoV-2 Infection Among Health Care Personnel, First Responders, and Other Essential and Frontline Workers - Eight U.S. Locations, December 2020-March 2021. MMWR Morb. Mortal. Wkly. Rep. 70, 495–500 (2021).

34. Andrejko, K. L. et al. Prevention of COVID-19 by mRNA-based vaccines within the general population of California. Clin. Infect. Dis. (2021) doi:10.1093/cid/ciab640.

35. Cabezas, C. et al. Associations of BNT162b2 vaccination with SARS-CoV-2 infection and hospital admission and death with covid-19 in nursing homes and healthcare workers in Catalonia: prospective cohort study. BMJ 374, n1868 (2021).

36. Self, W. H. et al. Comparative Effectiveness of Moderna, Pfizer-BioNTech, and Janssen (Johnson & Johnson) Vaccines in Preventing COVID-19 Hospitalizations Among Adults Without Immunocompromising Conditions - United States, March-August 2021. MMWR Morb. Mortal. Wkly. Rep. 70, 1337–1343 (2021).

37. Dickerman, B. A. et al. Comparative effectiveness of BNT162b2 and mRNA-1273 vaccines in U.s. veterans. N. Engl. J. Med. (2021) doi:10.1056/nejmoa2115463.

38. Payne, R. P. et al. Sustained T Cell Immunity, Protection and Boosting Using Extended Dosing Intervals of BNT162b2 mRNA Vaccine. (2021) doi:10.2139/ssrn.3891065.

39. Rosenberg, E. S. et al. Covid-19 Vaccine Effectiveness in New York State. N. Engl. J. Med. (2021) doi:10.1056/NEJMoa2116063.

40. Naranbhai, V. et al. Comparative immunogenicity and effectiveness of mRNA-1273, BNT162b2 and Ad26.COV2.S COVID-19 vaccines. J. Infect. Dis. (2021) doi:10.1093/infdis/jiab593.

41. NYC Department of Health and Mental Hygience. COVID-19 Data. New Variants/Strains. https://www1.nyc.gov/site/doh/covid/covid-19-data-variants.page (2021).

42. Fowlkes, A. et al. Effectiveness of COVID-19 Vaccines in Preventing SARS-CoV-2 Infection Among Frontline Workers Before and During B.1.617.2 (Delta) Variant Predominance – Eight U.S. Locations, December 2020-August 2021. MMWR Morb. Mortal. Wkly. Rep. 70, 1167–1169 (2021).

43. Gu, H. et al. Probable Transmission of SARS-CoV-2 Omicron Variant in Quarantine Hotel, Hong Kong, China, November 2021. Emerg. Infect. Dis. 28, (2021).

44. Stokes, E. K. et al. Coronavirus Disease 2019 Case Surveillance — United States, January 22– May 30, 2020. MMWR. Morbidity and Mortality Weekly Report vol. 69 759–765 (2020).

45. Rosenthal, N., Cao, Z., Gundrum, J., Sianis, J. & Safo, S. Risk Factors Associated With In-Hospital Mortality in a US National Sample of Patients With COVID-19. JAMA Netw Open 3, e2029058 (2020).

46. Price-Haywood, E. G., Burton, J., Fort, D. & Seoane, L. Hospitalization and Mortality among Black Patients and White Patients with Covid-19. N. Engl. J. Med. 382, 2534–2543 (2020).

47. Madhura S. Rane, Shivani Kochhar, Emily Poehlin, William You, McKaylee Robertson, Rebecca Zimba, Drew A. Westmoreland, Matthew L. Romo, Sarah Kulkarni, Mindy Chang, Amanda Berry, Christian Grov, and Denis Nash for the CHASING COVID Cohort Study Team. Determinants and trends of COVID-19 vaccine hesitancy and vaccine uptake in a national cohort of U.S. adults: A longitudinal study. Am. J. Epidemiol.

48. Nambi Ndugga, Latoya Hill, Samantha Artiga, Sweta Haldar. Latest Data on COVID-19 Vaccinations by Race/Ethnicity. (2022).

49. Shenai, M. B., Rahme, R. & Noorchashm, H. Equivalency of Protection From Natural Immunity in COVID-19 Recovered Versus Fully Vaccinated Persons: A Systematic and Pooled Analysis. Cureus 13, e19102 (2021).

50. Office for National Statistics (ONS). Coronavirus (COVID-19) Infection Survey Technical Article: Impact of vaccination on testing positive in the UK: October 2021. (2021).

51. Hall, V. J. et al. SARS-CoV-2 infection rates of antibody-positive compared with antibody-negative health-care workers in England: a large, multicentre, prospective cohort study (SIREN). Lancet 397, 1459–1469 (2021).

52. Gazit, S. et al. Comparing SARS-CoV-2 natural immunity to vaccine-induced immunity: reinfections versus breakthrough infections. bioRxiv (2021) doi:10.1101/2021.08.24.21262415.

53. Murugesan, M. et al. Protective Effect Conferred by Prior Infection and Vaccination on COVID-19 in a Healthcare Worker Cohort in South India. (2021) doi:10.2139/ssrn.3914633.

54. Bozio, C. H. et al. Laboratory-Confirmed COVID-19 Among Adults Hospitalized with COVID-19-Like Illness with Infection-Induced or mRNA Vaccine-Induced SARS-CoV-2 Immunity - Nine States, January-September 2021. MMWR Morb. Mortal. Wkly. Rep. 70, 1539–1544 (2021).

55. Haveri, A. et al. Persistence of neutralizing antibodies a year after SARS-CoV-2 infection. doi:10.1101/2021.07.13.21260426.

56. Alfego, D. et al. A population-based analysis of the longevity of SARS-CoV-2 antibody seropositivity in the United States. EClinicalMedicine 36, 100902 (2021).

57. Hansen, C. H., Michlmayr, D., Gubbels, S. M., Mølbak, K. & Ethelberg, S. Assessment of protection against reinfection with SARS-CoV-2 among 4 million PCR-tested individuals in Denmark in 2020: a population-level observational study. Lancet 397, 1204–1212 (2021).

58. Krammer, F. et al. Antibody Responses in Seropositive Persons after a Single Dose of SARS-CoV-2 mRNA Vaccine. N. Engl. J. Med. 384, 1372–1374 (2021).

59. Röltgen, K. et al. Defining the features and duration of antibody responses to SARS-CoV-2 infection associated with disease severity and outcome. Sci Immunol 5, (2020).

60. Wheatley, A. K. et al. Evolution of immune responses to SARS-CoV-2 in mild-moderate COVID-19. Nat. Commun. 12, 1162 (2021).

61. Canaday, D. H. et al. Significant reduction in humoral immunity among healthcare workers and nursing home residents 6 months after COVID-19 BNT162b2 mRNA vaccination. bioRxiv (2021) doi:10.1101/2021.08.15.21262067.

62. Bell, M. L. et al. Post-acute sequelae of COVID-19 in a non-hospitalized cohort: Results from the Arizona CoVHORT. PLoS One 16, e0254347 (2021).

63. Petersen, M. S. et al. Long COVID in the Faroe Islands: A Longitudinal Study Among Nonhospitalized Patients. Clin. Infect. Dis. 73, e4058–e4063 (2021).

64. Dean, N. E., Hogan, J. W. & Schnitzer, M. E. Covid-19 Vaccine Effectiveness and the Test-Negative Design. N. Engl. J. Med. (2021) doi:10.1056/NEJMe2113151.

65. Yan, Y. et al. Measuring voluntary and policy-induced social distancing behavior during the COVID-19 pandemic. doi:10.1101/2020.05.01.20087874.

66. Yuan, Y. et al. Changes in Mental Health and Preventive Behaviors before and after COVID-19 Vaccination: A Propensity Score Matching (PSM) Study. Vaccines (Basel) 9, (2021).

67. Kim, S. S. et al. mRNA Vaccine Effectiveness against COVID-19 among Symptomatic Outpatients Aged ≥16 Years in the United States, February – May 2021. doi:10.1101/2021.07.20.21260647.

68. Siegler, A. J. et al. Trajectory of COVID-19 Vaccine Hesitancy Over Time and Association of Initial Vaccine Hesitancy With Subsequent Vaccination. JAMA Netw Open 4, e2126882 (2021).

