## Supplementary for "Effectiveness of Covid-19 vaccines against symptomatic and asymptomatic SARS-CoV-2 infections in an urgent care setting"

Supplementary figure 1: Proportion of testers at CityMD by vaccination status between April 1, 2021 - October 25th, 2021

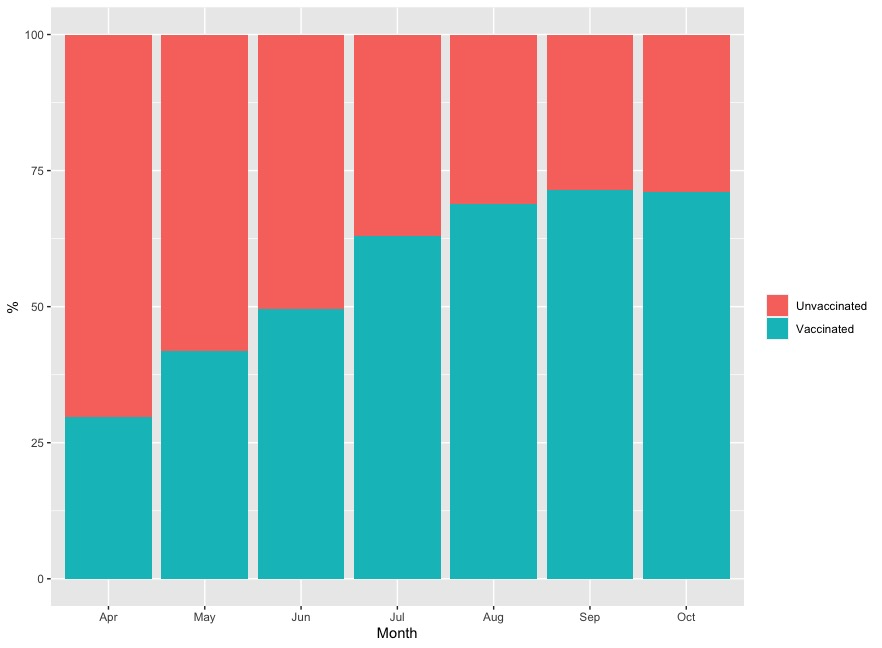

Supplementary table 1: Vaccine effectiveness against infections detected using RT-PCR only

|  | **Partial (Crude)** | | **Partial (Adjusted*)** | | **Fully (Crude)** | | **Fully (Adjusted)** | |
| --- | --- | --- | --- | --- | --- | --- | --- | --- |
|  | **VE** | **95% CI** | **VE** | **95% CI** | **VE** | **95% CI** | **VE** | **95% CI** |
| mRNA vaccines Pre-Delta | 0.50 | 0.47, 0.54 | 0.48 | 0.43, 0.52 | 0.91 | 0.89, 0.92 | 0.87 | 0.85, 0.89 |
| mRNA vaccines Post-Delta | 0.46 | 0.40, 0.51 | 0.48 | 0.42, 0.52 | 0.55 | 0.53, 0.57 | 0.67 | 0.59, 0.63 |
| Viral vector vaccine Pre-delta | -- | -- | -- | -- | 0.58 | 0.51, 0.65 | 0.46 | 0.35, 0.54 |
| Viral vector vaccine Post delta | -- | -- | -- | -- | 0.21 | 0.14, 0.28 | 0.28 | 0.21, 0.34 |

* Adjusted for age, gender, race, region, high exposure risk, comorbidities, calendar time (2 week periods)
